# Assessing Direct and Spillover Effects of Intervention Packages in Network-Randomized Studies

**DOI:** 10.1101/2022.03.24.22272909

**Authors:** Ashley L. Buchanan, Raúl Ulises Hernández-Ramírez, Judith J. Lok, Sten H. Vermund, Samuel R. Friedman, Laura Forastiere, Donna Spiegelman

**Author notes:** Contact for corresponding author (Buchanan): 7 Greenhouse Road, Kingston, RI 02881. DATA AND COMPUTING CODE AVAILABILITY: Example SAS code used to fit the models presented in this paper can be found in the Supplement Appendix E. The HPTN 037 study data sets are publicly available and can be requested from the Statistical Center for HIV/AIDS Research and Prevention through the ATLAS Science Portal through https://atlas.scharp.org/cpas.

## Abstract

Intervention packages may result in a greater public health impact than single interventions. Understanding the separate impact of each component in the overall package effectiveness can improve intervention delivery. We adapted an approach to evaluate the effects of a time-varying intervention package in a network-randomized study. In some network-randomized studies, only a subset of participants in exposed networks receive the intervention themselves. The spillover effect contrasts average potential outcomes if a person was not exposed themselves under intervention in the network versus no intervention in a control network. We estimated effects of components of the intervention package in HIV Prevention Trials Network 037, a Phase III network-randomized HIV prevention trial among people who inject drugs and their risk networks using Marginal Structural Models to adjust for time-varying confounding. The index participant in an intervention network received a peer education intervention initially at baseline, then boosters at 6 and 12 months. All participants were followed to ascertain HIV risk behaviors. There were 560 participants with at least one follow-up visit, 48% of whom were randomized to the intervention, and 1,598 participant-visits were observed. The spillover effect of the boosters in the presence of initial peer education training was a 39% rate reduction (Rate Ratio = 0.61; 95% confidence interval= 0.43, 0.87). These methods will be useful to evaluate intervention packages in studies with network features.

## Introduction

Design and scale-up of intervention packages could help to control the HIV/AIDS epidemic by meeting target population needs [1–3]. Intervention packages are defined herein as a set of individual components to simultaneously prevent or treat a disease or condition through multiple pathways. Many interventions confer partial protection against HIV transmission [1, 4]. Offering these interventions in combination (e.g., HIV testing, treatment as prevention (TasP)) is an important strategy. TasP is HIV treatment that suppresses an individual’s viremia and also prevents onward transmission [5]. Several cluster-randomized trials of packages of HIV prevention and treatment interventions were conducted in Sub-Saharan Africa and demonstrated a range of effectiveness for TasP in combination with other HIV interventions [6–13]. Intervention packages are often tailored to specific subpopulations, such as people who inject drugs (PWID), in an effort to achieve a larger and more sustainable intervention impact [14–16].

In studies of intervention packages, treatment may be randomly assigned to individuals and/or networks (e.g., social groupings), or treatments can be self-selected by individuals. Previous causal inference approaches accounted for implementation factors that were not randomized [17, 18], but did not consider possible spillover. Methods to evaluate the joint causal effects of two non-randomized exposures [19, 20] considered an *interaction* in the presence of time-varying confounding in an observational study, employing joint Marginal Structural Models (MSMs), but did not consider spillover effects. He et al. [21] developed MSMs for studies of a single intervention with spillover and fit the models using cluster-level propensity scores. MSMs are a class of causal models that typically model the marginal mean of the counterfactual outcome, and the parameters correspond to average causal effects [22].

We adapted methods to disentangle the effects of time-varying components of intervention packages in studies where spillover may be present. We considered a network-randomized trial in which only one member of each intervention network was eligible to be exposed to the HIV intervention package by the study investigators and each participant belongs to only one network. The participants eligible to receive the intervention package are hereafter denoted as “index participants”. Participants came forward to be an index, then centered around each index, investigators ascertained their immediate HIV risk contacts, defined as all individuals reporting injection or sexual behavior with the index, known as an *egocentric network*. These networks were randomized to intervention (index is peer educator) or control condition. After the initial training, the index participants in intervention networks then educated their network members (all participants in the network besides the index) about HIV risk reduction behavior. At 6and 12-month visits, the index participant could attend a voluntary booster session (second component) aimed at strengthening the initial intervention (first component). This study design is frequently utilized in HIV prevention research among PWID [23–26]. There was only one intervention component per visit; however, the MSM approach presented herein is more broadly applicable to multiple components per visit and aimed at evaluating joint causal effects. With only one component per visit, a time-varying mediation approach could alternatively be used with the exposure defined as the initial component and the subsequent components defined as mediators to understand how the initial intervention operates through the subsequent boosters [27].

Each component of the intervention package is considered a time-varying non-randomized exposure possibly subject to time-varying confounding. Effects can be estimated using inverse probability weights to fit MSMs [28, 29]. MSMs can also be used to estimate spillover effects when only one network member is exposed [30]. The *spillover* (indirect, disseminated) effect compares the risk of the outcome if a participant is a network member (non-index) comparing network exposure to package components versus no exposure [31, 32]. The *direct* effect compares the risk of the outcome if a participant is an index versus a network member under network exposure to package components. Illustrating with data from a network-randomized trial in the HIV Prevention Trials Network (HPTN) [33, 34], we employ inverse probability weighted (IPW) generalized linear mixed models to estimate these effects, adjusting for time-varying confounding.

## Methods

### Notation

Let uppercase letters denote random variables and lowercase letters denote realizations of those random variables. We define the network as the index participant and their egocentric network. Let *K* be the total number of networks and *i* = 1, …, *n*_*k*_ denote *n*_*k*_ participants in network *k*. Let ∑_*k*_ *n*_*k*_ = *N* be the total number of participants in the study, *j* = 0, …, *m*_*ki*_ denote the study visit for participant *i* in network *k*, where visit 0 corresponds to the baseline visit, *J*_*k*_ = ∑_*i*_ *m*_*ki*_, and *M*_*k*_ = max_*i*_ *m*_*ki*_. Define *Y*_*kij*_ as the observed binary outcome for participant *i* in network *k* at visit *j*. Let *X*_*k*_ be the randomized intervention package for network *k* and let *R*_*ki*_ denote index (versus network member) status for participant *i* in network *k*. Let *A*_*hkj*_ denote exposure to the *h*^*th*^ intervention component for network *k* at visit *j*, with *h* = 1, …, *H*. Let Ā _*hkj*_ = (*A*_*hk*0_, *A*_*hk*1_, …, *A*_*hkj*_)^*T*^ be a vector of size (*j* + 1) × 1 denoting the exposure histories for component *h* up to and including visit *j* for network *k*. Let **A**_*kj*_ = (*A*_1*kj*_, …, *A*_*Hkj*_)^*T*^ denote the vector of size *H* × 1 denoting all package component exposures in network *k* at visit *j* and let the history of the intervention package component exposures be denoted by the matrix

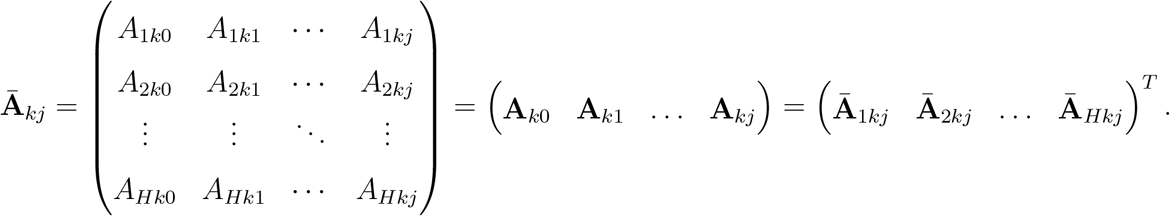

We assume that there is only one index per network and let *i*_*k*_ denote the unique index member in each network *k*. Let 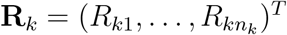 be a vector of size *n*_*k*_ × 1 denoting the index membership in network *k*. Let 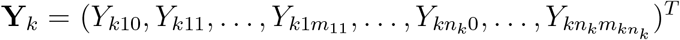 be the vector of size *J*_*k*_ × 1 denoting outcomes across all participant-visits in network *k*. Let **Z**_*ki*_ denote a *p* × 1 vector of baseline (i.e., time invariant) covariates for participant *i* in network *k* and **Z**_*kij*_ denote a *q* × 1 vector of time-varying covariates for participant *i* in network *k* at visit *j*. Let 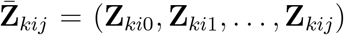 denote a *p* × *j* matrix for an individual’s covariate history up to and including visit *j*. At the network level, let 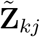 be a *qj* × 1 vector of network-level aggregate functions of the covariates, such as the history of the mean for a given covariate, in network *k* up to and including visit *j*. Let 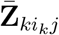 the index member covariates in network *k* up to and including visit *j*. Let **U**_*ki*_ be a vector of size *s* × 1 of unmeasured baseline covariates for participant *i* in network *k*. Let **U**_*k*_ denote a vector of the unmeasured baseline covariates of all participants in network *k*. Let *ā*_*j*_ denote a possible history of all package components from baseline up to and including visit *j* and ***r*** a realization of **R**_*k*_. Assuming partial interference (i.e., allowing for spillover between any members of an egocentric network but not between networks) [35], each participant has potential outcomes **Y**_*ki,j*+1_(***r***, *ā*_*j*_), which correspond to the 2^*H×j*^ × *n*_*k*_ vector of potential outcomes for participant *i* in network *k* at visit *j* under the index status indicator vector **R**_*k*_ = ***r*** and package component history Ā_*kj*_ = *ā*_*j*_. Assuming only one index per network and that the potential outcomes depend on the package component history and index status of the participant, but not specifically who is the index (i.e., stratified interference assumption [35]), the potential outcomes of interest are *Y*_*ki,j*+1_(*r, ā*_*j*_).

### Estimands

The *direct* package effect is a contrast in average potential outcomes under index versus network member status if the network is exposed component history *ā*_*j*_. On the ratio scale, *RR*^*D*^(ā_*j*_) = *E*[*Y*_*ki,j*+1_(*r* = 1, *ā*_*j*_)]*/E*[*Y*_*ki,j*+1_(*r* = 0, *ā*_*j*_)]. The *spillover* package effect compares the average potential outcomes if a participant is a network member under network component history *ā*_*j*_ versus no component history exposure 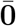. 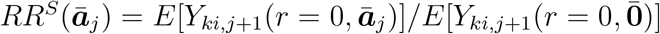. The *composite* package effect is 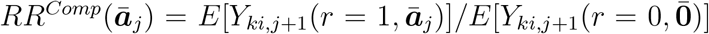, a comparison of the average potential outcomes if an index under network component history exposure *ā*_*j*_ versus if a network member under no component history exposure 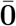. Marginalizing over the index status, the *overall* package effect compares average potential outcomes under component history exposure *ā*_*j*_ versus no exposure, denoted 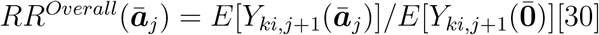 Analogous effects can be defined on the difference scale and other definitions of these effects are possible.

### Assumptions

We assume there may be correlation between outcomes in a network and no contamination across study intervention arms (e.g., the intervention does not affect participants in the control group) [31]. At the individual level conditional on **Z**_*ki*_, we assume conditional *index member exchangeability* for the self-selected index status; that is, *Y*_*ki,j*+1_(*r, ā*_*j*_) ⊥ **R**_*ki*_|**Z**_*ki*_ [30, 36]. *For the time-varying package components* ***A***_*kj*_, *network component exchangeability* may not hold because the network-level exposure is determined by index visit attendance that may depend on covariates. However, we assume 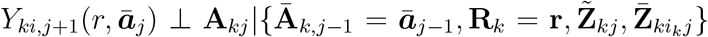 for *j* = 0, …, *m*_*ki*_; that is, network component exchangeability of potential outcomes holds conditional on each network’s package component history (determined by the network-level intervention, index status, and index visit attendance) and a network-level aggregate of covariate history, as well as its index member’s covariate history, up to and including the prior visit *j*. In the HPTN trial, covariates are ascertained from previous visits because the component exposure is defined by index visit attendance, which can only be modeled across all indexes using information from previous visits. We assume positivity: 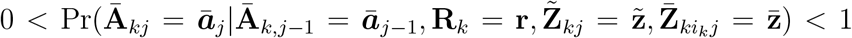 for all *r, ā*_*j*_, **r, *z***, 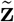 and 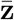, requiring index and non-index members at each level of individual covariates and networks exposed (and not exposed) to each of the components at each level of the index and network-level covariates. The latter can hold when there is non-compliance to package components. If two components were always delivered simultaneously, we would not be able to disentangle the package component effects. Related to causal consistency, we also assume treatment variation irrelevance for the package components, which implies we have only one version of the outcome for exposure to each component and one version for no exposure [37]. We also assume that data from missed visits and dropout due to loss-to-follow-up are Missing Completely at Random [38].

## Estimation and Inference

We estimate the effect of each package component in a single model, while considering the presence of the remaining package components [30]. This outcome model is adjusted for individual-level confounding at baseline and IPW to adjust for network level time-varying confounding (Appendix A). We assume that the effects of the package components and index status are not modified by covariates (e.g., there are no (*r, *ā**_*j*_) by **Z**_*ki*_ interactions) and the log-binomial generalized linear mixed model fits the data. The MSM is

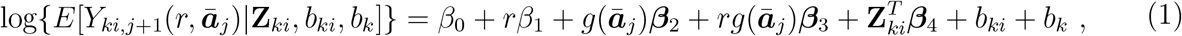

where *g*(ā_*j*_) is a known function of the package component history with 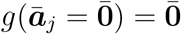 and the param-eters ***β***_2_, ***β***_3_ are column vectors with dimensions determined by *g*(ā_*j*_). For example, 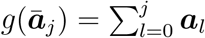 could be defined as cumulative exposure or a simple time-updated exposure with exposure status at baseline for each package component, 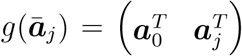. Two random intercepts, *b*_*ki*_ ∼ *N* (0, *ψ*_1_) and *b*_*k*_ ∼ *N* (0, *ψ*_2_), account for correlation within participants and within networks, respectively. We assume the corresponding observed outcomes 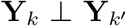 are independent for *k* ≠ *k*^*′*^ . The vector of unmeasured baseline covariates **U**_*ki*_ may affect *R*_*ki*_ within levels of the baseline covariates **Z**_*ki*_. The *β*_1_ term represents outcome differences between index and non-index members in the absence of the intervention after conditioning on **Z**_*ki*_. Note that 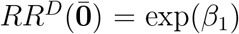. We assume that index status can only have an effect on the outcome in an intervention network because an index in an intervention network is the only individual assigned to receive the training from the study team (i.e., peer educator); that is, 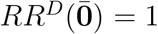 and thus *β*_1_ = 0. If conditional exchangeability does not hold for index status, 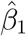 could be different from the null due to unmeasured differences between index participants and network members. The differences in the observed outcomes between index participants in the intervention arm and network members in the intervention arm (for direct effect) or in the control arm (for the composite effect) are due to both the causal effects and the differences between the two groups due to unmeasured confounding. Therefore, dividing by its exponent exp(*β*_1_) on the ratio scale would result in the causal effects. To note, identification of the MSM parameters under unmeasured confounding of index status is a consequence of the functional form of the model. Extending results in Buchanan et al. [30], suppressing the notation for the random effects and covariates for ease of notation, estimators for each of the parameters are:

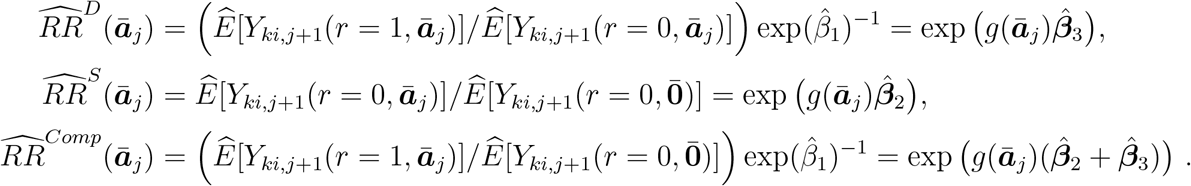

On the relative rate scale under the assumptions described above, *RR*^*D*^(ā) can be interpreted as the causal spillover effect if a participant is a network member under network exposure to intervention package component history exposure *ā*_*j*_ versus no network exposure. For log links, the conditional mean is additive in the fixed and random effects rendering the conditional and marginal treatment effects equivalent and these estimated effects can be interpreted as either participant-level and/or population-level estimates [39, 40]. We consider a model for quantifying the effects of multiple package components that includes pairwise interactions between components in a single model (Appendix B). Ignoring the estimation of the weights [41], we used a conservative empirical sandwich estimator of the variance from a generalized linear mixed model with a working binomial variance and independent correlation specified by within-participant and within-network random intercepts to construct 95% Wald-type confidence intervals (CIs) [38, 42].

## Illustrative Example

The HPTN Protocol 037 was a Phase III, network-randomized controlled HIV prevention trial with 696 participants who were PWID and their HIV risk networks in Philadelphia, PA [14]. HPTN 037 evaluated the efficacy of a network-oriented peer education intervention package to promote HIV risk reduction among HIV risk networks. Participants were followed for up to 30 months with visits every six months. Index participants whose network was randomized to the intervention received an intervention package that consisted of an initial intervention at baseline with six 2hour peer educator sessions during the first four weeks and boosters at six and 12 months. The initial training educated indexes in the intervention networks through role-playing activities and developing communication skills including: 1) promote safer sex and drug injection skills among network members, and 2) communication strategies to reach peers and promote norms about HIV risk reduction. At the booster sessions, indexes had the opportunity to troubleshoot, share their experiences, improve communication, and bolster motivation. An intervention network was exposed if its index member attended a visit when a package component (initial or boosters) was administered. Participants in both the intervention and control conditions received HIV counseling and testing at each visit [14]. Table 1 presents the intervention package evaluated in HPTN 037.

**Table 1:** Interventions in HPTN 037 by study arm and network membership.

| Study Arm | Behavioral Intervention |  |
| --- | --- | --- |
|  | Index Participant | Network Members |
| **Experimental Arm** | Enhanced HIV counseling and testing *plus* Six 2-hour network-orientated peer-educator sessions during weeks 1 to 4 (initial intervention) *and* Booster session at months 6 and 12 | Enhanced HIV Counseling and Testing |
| **Control Arm** | Enhanced HIV Counseling and Testing | Enhanced HIV Counseling and Testing |

We are interested in the overall effect of the additional boosters among those who received the initial intervention and the direct and spillover effects of the intervention package components (i.e, initial and boosters). Exposure to the booster was defined as a time-updated variable for the booster exposure at time *j* with the outcome at time *j* + 1. The package components were defined as initial only and recent exposure to the 6or 12-month booster in addition to the initial intervention. Across all study visits, we estimated effects of the initial intervention only and the boosters in the presence of the initial intervention that describe the patterns of change in the response probability over time (Appendix D). We evaluated the effects at the 6-, 12-, and 18-month visits (following the delivery of the initial intervention) and 12and 18-month visits (following the delivery of each booster intervention). Direct and spillover effects of the package components were assessed through comparisons of the report of any injection-related risk behavior across all study visits defined by: sharing injection equipment (needles, cookers, cotton, and rinse water), front and back loading (i.e., injecting drugs from one syringe to another), and injecting with people not well known or in a shooting gallery/public place in the past month. Following the original analysis of this study [14], outcomes among participants who reported injection drug use in the six months prior to baseline were included. The longitudinal data were used to assess the effects of the intervention on the inter-visit rates of any injection-related risk behavior using a multilevel generalized linear mixed model (see Estimation and Inference).

Ideally, we would include all covariates that were known or suspected risk factors; however, this resulted in overparameterization of the model given the study’s sample size. We employed a variable selection procedure (Appendix C). The outcome models were adjusted for baseline individual-level covariates that were known or suspected risk factors for the outcome, including race (nonwhite vs. white), ethnicity (Hispanic vs. non-Hispanic), report of any injection risk behavior (yes vs. no), injected daily in the last month (yes vs. no), alcohol use (got drunk vs. no), and injected heroin and cocaine (yes vs. no) and network-level average age and prevalence of nonwhite race, report of any injection risk behavior, cocaine use, and injected heroin and cocaine. Models with time-varying covariates adjusted for the same set of individual-, index-, and network-level covariates included in the baseline model, with baseline index- and network-level covariates replaced with their time-varying version when applicable.

Time-varying confounding was adjusted for using stabilized inverse probability weights (Appendix A). The IPW generalized linear mixed model with a log link and binomial distribution was fit by weighting individual participants according to their estimated stabilized weights. In intervention networks only, the weight models were estimated in the network-level data with one record per network per visit, pooling across the visits where the network-level exposure status could change (e.g., 12to 18-month visits). The denominator weight models included selected time-varying networklevel aggregate covariates, index member covariates that were known or suspected risk factors for the outcome, and selected pairwise interactions, which allowed for more flexibility in the model specification, and the package component exposure at the previous visit. The numerator weight model included the package component exposure at the previous visit and selected baseline network-level covariates and index member covariates that were known or suspected risk factors for the outcome, which were also included in the outcome model. Both the models for the numerator and denominator of the weights included a variable for time specified as study visit month. For some of the models, the log-binomial models did not converge and log-Poisson models, which provide consistent but not fully efficient estimates of the relative risk, were used [39, 43]. There was no evidence of model misspecification or positivity violations because the average of the stabilized weights distribution was approximately one (mean = 0.99; standard deviation = 0.08; minimum = 0.10; maximum = 2.13; Table A1), using the criterion in [44]. For comparison to traditional methods, we presented models adjusting for baseline covariates only or including the time-varying covariates in the outcome model, and both methods may result in estimates that do not confer a causal interpretation due to bias (Appendix A). Additional results, including a sensitivity analysis for effect measure modification by study visit month, are provided (Appendix D, Tables A2 and A3). The data analysis for this paper was generated using SAS software (Version 9.4), and we provide example SAS code (Appendix E).

## Results

There were 696 participants, 651 reported injection drug use at baseline, and 560 participants had at least one follow-up visit with a total of 1,598 person visits. The size of the networks ranged from two to seven participants and network size was not associated with the outcome (unadjusted rate ratio (RR) = 0.96; 95% CI = 0.87, 1.06). Among the 560 participants, 270 (48%) were in intervention networks. Of the 232 indexes who reported injection drug behavior at baseline, 112 (48%) received the initial peer education intervention. Table 2 displays the number of participants who received initial peer education and each booster session; in intervention networks, 70 (63%) index participants received the 6-month booster, and 59 (53%) index participants received the 12-month booster.

**Table 2:** Package components received by index and network members during follow-up among HPTN 037 participants who reported injecting drugs at baseline and had at least one follow-up visit after baseline (n = 560)

|  | **Network Member Role** |  |
| --- | --- | --- |
|  | **Index** | **Network Member<sup>1</sup>** |
| Initial Peer Education ($n = 560$)<sup>2</sup> | 97/201 (48%) | 173/359 (48%) |
| 6-Month Booster ($n = 447$) | 70/159 (44%) | 128/288 (44%) |
| 12-Month Booster ($n = 344$) | 59/125 (47%) | 106/219 (48%) |

<sup>1</sup>Network member exposure determined by randomized package and their index's visit attendance.
<sup>2</sup> Reported $n$ is the total number of participants who had at least some follow-up after baseline.

Regardless of the adjustment approach, the estimated spillover effect for the initial peer education intervention was comparable (Table 3). In the IPW models, there was an estimated 13% rate reduction in the spillover rate of report of any injection risk behavior (RR = 0.87; 95% CI = 0.69, 1.10). We expect a spillover rate of reporting any injection risk behavior to be 39% lower if a participant is a network member in an intervention network whose index had recent exposure to the 6or 12-month booster in addition to the initial training, compared to if a participant is a control network member (RR = 0.61; 95% CI = 0.43, 0.87). The estimated direct effect of the initial peer education intervention was protective, regardless of the covariate adjustment approach; however, the estimated direct effect of the booster in addition to the initial intervention was null. In the IPW models, the estimated initial and booster composite effects were protective with a 24% (RR = 0.76; 95% CI = 0.56, 1.02) and 37% rate reduction (RR = 0.63; 95% CI = 0.43, 0.92), respectively. The estimated overall effect of the initial was protective with a 17% rate reduction (RR = 0.83; 95% CI = 0.69, 0.99). We expect a 38% decrease in the overall rate of reporting injection risk behavior if networks are recently exposed to the 6or 12-month booster in addition to the initial training compared to if networks were assigned to the control condition (RR = 0.62; 95% CI = 0.46, 0.82). Based on the MSM parameterization, the rate ratios could be interpreted as the estimates from a trial in which participants are randomized to the booster at six months, 12 months, or no booster at each visit [45]. The effect measure modification of the four different intervention effects by visit time on the multiplicative scale are reported in Table A3. The magnitude of the estimated spillover effect of the booster was larger at 12 months (RR = 0.54; 95% CI = (0.36, 0.81)), compared to 18 months (RR = 0.69; 95% CI = 0.44, 1.10; P value = 0.54). The magnitude of the estimated spillover effect of the initial intervention was comparable across study visits (P value = 0.96).

**Table 3:** Estimated rate ratios (RR) for the direct, spillover, composite and overall effects of the HPTN 037 peer education package components on reducing report of any injection risk behavior (per person-visit) during follow-up with 95% confidence intervals (CI) among participants with at least one follow-up visit^1^

|  | Direct |  | Spillover |  |
| --- | --- | --- | --- | --- |
|  | Initial | Booster | Initial | Booster |
| Unadjusted | 0.94 (0.65, 1.36) | 1.03 (0.64, 1.66) | 0.86 (0.66, 1.11) | 0.59 (0.39, 0.87) |
| Adjusted for baseline covariates<sup>2</sup> | 0.87 (0.59, 1.26) | 1.05 (0.65, 1.68) | 0.87 (0.68, 1.10) | 0.62 (0.43, 0.88) |
| Adjusted for baseline and TV covariates<sup>2,3</sup> | 0.83 (0.57, 1.21) | 1.00 (0.62, 1.64) | 0.88 (0.69, 1.11) | 0.72 (0.49, 1.06) |
| IP-weighted<sup>2,4</sup> | 0.87 (0.60, 1.27) | 1.03 (0.64, 1.65) | 0.87 (0.69, 1.10) | 0.61 (0.43, 0.87) |
|  | Overall |  | Composite |  |
|  | Initial | Booster | Initial | Booster |
| Unadjusted | 0.84 (0.68, 1.04) | 0.59 (0.43, 0.82) | 0.80 (0.59, 1.10) | 0.60 (0.41, 0.89) |
| Adjusted for baseline covariates<sup>2</sup> | 0.82 (0.68, 0.99) | 0.63 (0.47, 0.83) | 0.75 (0.56, 1.01) | 0.64 (0.44, 0.94) |
| Adjusted for baseline and TV covariates<sup>2,3</sup> | 0.82 (0.68, 0.98) | 0.72 (0.54, 0.96) | 0.73 (0.55, 0.96) | 0.72 (0.51, 1.02) |
| IP-weighted<sup>2,4</sup> | 0.83 (0.69, 0.99) | 0.62 (0.46, 0.82) | 0.76 (0.56, 1.02) | 0.63 (0.43, 0.92) |

<sup>1</sup> Analysis included a total of 560 participants and 1,598 person-visits with 509 events total. One participant was excluded due to missing baseline information on spending the night on the street (in the past 6 months) and spending time in jail (in the past 6 months) at baseline. The package components were defined as initial only and recent exposure for the 6- or 12-month booster in addition to the initial intervention.
<sup>2</sup> Baseline covariates included individual-level race (nonwhite vs. white), Hispanic (yes vs. no), report of any injection risk behavior (yes vs. no), injected daily in the last month (yes vs. no), and alcohol use (got drunk vs. no), index member race (nonwhite vs. white), index report of any injection risk behavior (yes vs. no), index injected daily in the last month (yes vs. no), and index injected heroin and cocaine (yes vs. no) and network-level average age and network-level prevalence of nonwhite race, report of any injection risk behavior, cocaine use, and injected heroin and cocaine.
<sup>3</sup> Adjusted for the same individual-level covariates included in the baseline model, network-level prevalence of report of any injection risk behavior at baseline, index member race, baseline network-level average age, baseline network-level prevalence of nonwhite race, and also the time-varying version of the remaining index member and network-level covariates included in the baseline model, except for time-varying index member report of injected heroin and cocaine (Appendix C).
<sup>4</sup> Adjusted for weights estimated using the same index member and network-level covariates included in the baseline model, time-varying network-level prevalence of report of any injection risk behavior, cocaine use, and injected heroin and cocaine, the time-varying index report of any injection risk behavior and index injected daily in the last month, and four interaction terms between selected time-varying covariates: index report of any injection risk behavior and injected daily in the last month; network-level prevalence of injected heroin and cocaine and index report of any injection risk behavior; network-level prevalence of injected heroin and cocaine and index injected daily in the last month; and network-level prevalence of injected heroin and cocaine and report of any injection risk behavior.

## Discussion

We adapted existing causal inference methods to evaluate time-varying components of intervention packages in studies where spillover may be present. Estimating these effects provides information about the impact of package interventions and their spillover to network members from an exposed index participant. These methods provide a more in-depth understanding of package components’ effects for exposed index participants and those sharing networks with exposed participants. In HPTN 037, the estimated overall effect was larger in magnitude for the booster with initial intervention, as compared to the initial intervention alone, highlighting the importance of continued training for peer educators in this context. A protective spillover effect was observed for network members for the initial and booster interventions, without a corresponding direct benefit for those trained to be peer educators. We found evidence that booster sessions strengthen the spillover of the intervention, which can be utilized when developing peer-led interventions [46].

This work is particularly timely as these methods are applicable to several HIV combination prevention cluster-randomized trials that warrant evaluation of the intervention package component effects with multiple components at each visit, as well as the spillover effects within communities, to better understand the effects of TasP [3, 47]. Understanding the components that may be driving the observed effects of the intervention package could inform modifications to the existing public health strategies in these settings by strengthening highly effective components and redeveloping those found to be partially effective or ineffective [6–8, 10, 12, 13]. Evaluation of the universal “test and treat” intervention could include the impact on the health outcomes among those in the communities assigned to immediate ART scale-up but who did not receive immediate ART themselves. As in vaccine campaign design, this could inform the level of ART coverage needed to benefit the community and consistently achieve targets such as the UNAIDS 2025 [48]. Potential impact of a number of other HIV prevention strategies could be informed by estimating spillover effects in the community or social network [49–51].

The assumption of no unmeasured covariates associated with the network-level exposure and outcome is untestable. Future work could develop methods to assess the sensitivity of these methods to unmeasured confounders [52]. We assumed that index status has no effect in the absence of an intervention conditional on measured confounders; however, if features of the intervention are not exclusive to the study and the indexes are more (or less) health seeking than others in their network, there could be a non-null index effect in the control. The weight and outcome models were assumed to be correct (e.g., correct functional forms of covariates). This is not guaranteed in practice, and sensitivity analyses could be performed to evaluate the robustness of results to model specification. We do not expect any structural violations of positivity, as both the index status and each of the package components can occur at each level of the measured covariates; however, given the number of covariates included, we used a parametric model for the weights [44]. We also assumed that any data from missing visits were ignorable with respect to valid estimation of intervention effects. These models could be extended to include censoring weights to adjust for possibly differential loss to followup due to dropout [53, 54]. The ascertainment of personal networks of index participants is likely only a partial ascertainment of each person’s network, which limits the evaluation of spillover within the networks to recruited individuals. These methods will be a valuable tool to evaluate randomized and non-randomized intervention packages in a single study with network features.

## Supporting information

Supplemental Files Revised

## Data Availability

Example SAS code used to fit the models presented in this paper can be found in the Supplement Appendix E. The HPTN 037 study data sets are publicly available and can be requested from the Statistical Center for HIV/AIDS Research and Prevention through the ATLAS Science Portal through https://atlas.scharp.org/cpas/.

https://atlas.scharp.org/cpas/project/HPTN/

## ABBREVIATIONS

(CI): Confidence interval
(GEEs): Generalized estimating equations
(HPTN): HIV Prevention Trials Network
(HIV): Human Immunodeficiency Virus
(PWID): People who inject drugs
(RR): Risk/Rate ratio

## Acknowledgements

We thank Drs. Deborah Donnell and Carl Latkin for access to the HIV Prevention Trials Network (HPTN) 037 data and their valuable suggestions. Data from the HPTN were obtained with support from the National Institute on Drug Abuse (NIDA), National Institute of Allergies and Infectious Disease (NIAID), and National Institute of Mental Health (NIMH) under National Institutes of Health (NIH) grants UM1AI068x619 (HPTN Leadership and Operations Center), UM1AI068617 (HPTN Statistical and Data Management Center), and UM1AI068613 (HPTN Laboratory Center).

## References

1. Padian NS, McCoy SI, Manian S, Wilson D, Schwartländer B, Bertozzi SM. Evaluation of large-scale combination HIV prevention programs: essential issues. Journal of Acquired Immune Deficiency Syndromes. 2011; 58(2):e23–e28.

2. DeGruttola V, Smith DM, Little SJ, Miller V. Developing and evaluating comprehensive HIV infection control strategies: Issues and challenges. Clinical Infectious Diseases. 2010; 50(3):S102– S107.

3. Kurth AE, Celum C, Baeten JM, Vermund SH, Wasserheit JN. Combination HIV prevention: Significance, challenges, and opportunities. Current HIV/AIDS Reports. 2011; 8(1):62–72.

4. Padian NS, Holmes CB, McCoy SI, Lyerla R, Bouey PD, Goosby EP. Implementation science for the US President’s Emergency Plan for AIDS Relief (PEPFAR). Journal of Acquired Immune Deficiency Syndromes. 2011; 56(3):199–203.

5. Cohen MS, McCauley M, Gamble TR. HIV treatment as prevention and HPTN 052. Current Opinion in HIV and AIDS. 2012; 7(2):99–105.

6. Tanser F, Bärnighausen T, Grapsa E, Zaidi J, Newell ML. High coverage of ART associated with decline in risk of HIV acquisition in rural KwaZulu-Natal, South Africa. Science. 2013; 339(6122):966–971.

7. Hayes R, Moulton L. Cluster Randomised Trials. London: Taylor & Francis, 2009.

8. Hayes R, Ayles H, Beyers N, Sabapathy K, Floyd S, Shanaube K, et al. HPTN 071 (PopART): Rationale and design of a cluster-randomised trial of the population impact of an HIV combination prevention intervention including universal testing and treatment–a study protocol for a cluster randomised trial. Trials. 2014; 15(1):57.

9. Hayes RJ, Donnell D, Floyd S, Mandla N, Bwalya J, Sabapathy K, et al. Effect of universal testing and treatment on HIV incidence – HPTN 071 (PopART). New England Journal of Medicine. 2019; 381(3):207–218.

10. Iwuji CC, Orne-Gliemann J, Tanser F, Boyer S, Lessells RJ, Lert F, et al. Evaluation of the impact of immediate versus WHO recommendations-guided antiretroviral therapy initiation on HIV incidence: The ANRS 12249 TasP (Treatment as Prevention) trial in Hlabisa sub-district, KwaZulu-Natal, South Africa: Study protocol for a cluster randomised controlled trial. Trials. 2013; 14(1):230.

11. Iwuji CC, Orne-Gliemann J, Larmarange J, Balestre E, Thiebaut R, Tanser F, et al. Universal test and treat and the HIV epidemic in rural South Africa: A phase 4, open-label, community cluster randomised trial. The Lancet HIV. 2018; 5(3):e116–e125.

12. Chamie G, Kwarisiima D, Clark TD, Kabami J, Jain V, Geng E, et al. Leveraging rapid community-based HIV testing campaigns for non-communicable diseases in rural Uganda. PloS one. 2012; 7(8):e43400.

13. Chamie G, Kwarisiima D, Clark TD, Kabami J, Jain V, Geng E, et al. Uptake of communitybased HIV testing during a multi-disease health campaign in rural Uganda. PloS one. 2014; 9(1):e84317.

14. Latkin CA, Donnell D, Metzger D, Sherman S, Aramrattna A, Davis-Vogel A, et al. The efficacy of a network intervention to reduce HIV risk behaviors among drug users and risk partners in Chiang Mai, Thailand and Philadelphia, USA. Social Science and Medicine. 2009; 68(4):740– 748.

15. Nikolopoulos GK, Pavlitina E, Muth SQ, Schneider J, Psichogiou M, Williams LD, et al. A network intervention that locates and intervenes with recently HIV-infected persons: The Transmission Reduction Intervention Project (TRIP). Scientific Reports. 2016; 6.

16. Spoth R, Guyll M, Lillehoj CJ, Redmond C, Greenberg M. Prosper study of evidence-based intervention implementation quality by community–university partnerships. Journal of Community Psychology. 2007; 35(8):981–999.

17. Crowley DM, Coffman DL, Feinberg ME, Greenberg MT, Spoth RL. Evaluating the impact of implementation factors on family-based prevention programming: Methods for strengthening causal inference. Prevention Science. 2014; 15(2):246–255.

18. Lippman SA, Shade SB, Hubbard AE. Inverse probability weighting in STI/HIV prevention research: methods for evaluating social and community interventions. Sexually Transmitted Diseases. 2010; 37(8):512–518.

19. Hernán MA, Brumback B, Robins JM. Marginal structural models to estimate the joint causal effect of nonrandomized treatments. Journal of the American Statistical Association. 2001; 96(454):440–448.

20. Howe CJ, Cole SR, Mehta SH, Kirk GD. Estimating the effects of multiple time-varying exposures using joint marginal structural models: alcohol consumption, injection drug use, and HIV acquisition. Epidemiology. 2012; 23(4):574–582.

21. He J, Stephens-Shields A, Joffe M. Marginal structural models to estimate the effects of timevarying treatments on clustered outcomes in the presence of interference. Statistical Methods in Medical Research. 2019; 28(2):613–625.

22. Hernán MA, Robins JM. Causal Inference: What If. Boca Raton: Chapman & Hall/CRC, 2020.

23. Friedman SR, Bolyard M, Khan M, Maslow C, Sandoval M, Mateu-Gelabert P, et al. Group sex events and HIV/STI risk in an urban network. Journal of Acquired Immune Deficiency Syndromes. 2008; 49(4):440–446.

24. Khan MR, Epperson MW, Mateu-Gelabert P, Bolyard M, Sandoval M, Friedman SR. Incarceration, sex with an STI-or HIV-infected partner, and infection with an STI or HIV in Bushwick, Brooklyn, NY: A social network perspective. American Journal of Public Health. 2011; 101(6):1110–1117.

25. Friedman S, Bolyard M, Sandoval M, Mateu-Gelabert P, Maslow C, Zenilman J. Relative prevalence of different sexually transmitted infections in HIV-discordant sexual partnerships: Data from a risk network study in a high-risk New York neighbourhood. Sexually Transmitted Infections. 2008; 84(1):17–18.

26. Tobin KE, Kuramoto SJ, Davey-Rothwell MA, Latkin CA. The STEP into Action study: A peer-based, personal risk network-focused HIV prevention intervention with injection drug users in Baltimore, Maryland. Addiction. 2011; 106(2):366–375.

27. VanderWeele TJ, Tchetgen Tchetgen EJ. Mediation analysis with time varying exposures and mediators. Journal of the Royal Statistical Society Series B: Statistical Methodology. 2017; 79(3):917–938.

28. Hernán MA, Brumback B, Robins JM. Marginal structural models to estimate the causal effect of zidovudine on the survival of HIV-positive men. Epidemiology. 2000; 11(5):561–571.

29. Robins JM, Hernan MA, Brumback B. Marginal structural models and causal inference in epidemiology. Epidemiology. 2000; 11(5):550–560.

30. Buchanan A, Vermund S, Friedman S, Spiegelman D. Assessing Individual and Disseminated Effects in Network-Randomized Studies. American Journal of Epidemiology. 2018; 187(11):2449– 2459.

31. Benjamin-Chung J, Arnold BF, Berger D, Luby SP, Miguel E, Colford Jr JM, et al. Spillover effects in epidemiology: parameters, study designs and methodological considerations. International Journal of Epidemiology. 2018; 47(1):332–347.

32. Halloran ME, Hudgens MG. Dependent happenings: A recent methodological review. Current Epidemiology Reports. 2016; 3(4):297–305.

33. Sista ND, Karim QA, Hinson K, Donnell D, Eshleman SH, Vermund SH. Experience in international clinical research: the HIV Prevention Trials Network. Clinical Investigation. 2011; 1(124):1609–1618.

34. Vermund SH, Hodder SL, Justman JE, Koblin BA, Mastro TD, Mayer KH, et al. Addressing research priorities for prevention of HIV infection in the United States. Clinical Infectious Disease. 2010; 50 Suppl 3(Suppl 3):S149–55.

35. Sobel ME. What do randomized studies of housing mobility demonstrate? Causal inference in the face of interference. Journal of the American Statistical Association. 2006; 101(476):1398– 1407.

36. Ogburn EL, VanderWeele TJ. Causal diagrams for interference. Statistical Science. 2014; 29(4):559– 578.

37. VanderWeele TJ. Concerning the consistency assumption in causal inference. Epidemiology. 2009; 20(6):880–883.

38. Fitzmaurice GM, Laird NM, Ware JH. Applied Longitudinal Analysis. Hoboken: John Wiley & Sons, 2012.

39. Spiegelman D, Hertzmark E. Easy SAS calculations for risk or prevalence ratios and differences. American Journal of Epidemiology. 2005; 162(3):199–200.

40. Ritz J, Spiegelman D. Equivalence of conditional and marginal regression models for clustered and longitudinal data. Statistical Methods in Medical Research. 2004; 13(4):309–323.

41. Hernán MA, Brumback BA, Robins JM. Estimating the causal effect of zidovudine on CD4 count with a marginal structural model for repeated measures. Statistics in Medicine. 2002; 21 (12):1689–1709.

42. Stefanski LA, Boos DD. The calculus of M-estimation. The American Statistician. 2002; 56(1):29– 38.

43. Zou G. A modified poisson regression approach to prospective studies with binary data. American Journal of Epidemiology. 2004; 159(7):702–706.

44. Cole SR, Hernán MA. Constructing inverse probability weights for marginal structural models. American Journal of Epidemiology. 2008; 168(6):656–664.

45. Breskin A, Cole SR, Westreich D. Exploring the subtleties of inverse probability weighting and marginal structural models. Epidemiology. 2018; 29(3):352–355.

46. Lancaster KE, Miller WC, Kiriazova T, Sarasvita R, Bui Q, Ha TV, et al. Designing an individually tailored multilevel intervention to increase engagement in HIV and substance use treatment among people who inject drugs with HIV: HPTN 074. AIDS Education and Prevention. 2019; 31(2):95–110.

47. Perriat D, Balzer L, Hayes R, Lockman S, Walsh F, Ayles H, et al. Comparative assessment of five trials of universal HIV testing and treatment in sub-Saharan Africa. Journal of the International AIDS Society. 2018; 21(1):e25048.

48. De Lay PR, Benzaken A, Karim QA, Aliyu S, Amole C, Ayala G, et al. Ending AIDS as a public health threat by 2030: Time to reset targets for 2025. PLoS Medicine. 2021; 18(6):e1003649.

49. Brault MA, Spiegelman D, Hargreaves J, Nash D, Vermund SH. Treatment as prevention: Concepts and challenges for reducing HIV incidence. Journal of Acquired Immune Deficiency Syndromes. 2019; 82 Suppl 2(2):S104–S112.

50. Lou J, Hu P, Qian HZ, Ruan Y, Jin Z, Xing H, et al. Expanded antiretroviral treatment, sexual networks, and condom use: Treatment as prevention unlikely to succeed without partner reduction among men who have sex with men in China. PLoS One. 2017; 12(4):e0171295.

51. Zhang C, Webb GF, Lou J, Shepherd BE, Qian HZ, Li Y, et al. Predicting the long-term impact of voluntary medical male circumcision on HIV incidence among men who have sex with men in Beijing, China. AIDS Care. 2020; 32(3):343–353.

52. VanderWeele TJ, Tchetgen Tchetgen EJ, Halloran ME. Interference and sensitivity analysis. Statistical Science. 2014; 29(4):687–706.

53. Hernández-Ramírez RU, Spiegelman D, Lok JJ, Forastiere L, Friedman SR, Latkin CA, et al. Overall, direct, spillover, and composite effects of components of a peer-driven intervention package on injection risk behavior among people who inject drugs in the HPTN 037 study. AIDS and Behavior. 2024; 28(1):225–237.

54. Robins JM, Finkelstein DM. Correcting for noncompliance and dependent censoring in an AIDS clinical trial with inverse probability of censoring weighted log-rank tests. Biometrics. 2000; 56(3):779–788.

