## Supplemental Files Revised for "Assessing Direct and Spillover Effects of Intervention Packages in Network-Randomized Studies"

**The file includes:**

Appendix A: Development of models for weights to control for treatment confounder feedback

Appendix B: Approach for multiple package components with pairwise interactions

Appendix C: Covariate selection procedure for modeling in HPTN 037

Appendix D: Additional Results in HPTN 037

Appendix E: Example SAS Code

Table A1

Table A2

Table A3

References

### Appendix A

#### Development of models for weights to control for treatment confounder feedback

To guide our causal inference approach, an ideal experiment in this setting is conceptualized as a two-stage randomized design, where first networks are randomized to intervention or control, then individuals in each intervention network are randomized to be an index (or not) [1]. For an egocentric network randomized design, a single index would need to be randomly selected in each network. However, in practice, index assignment is typically not randomized, but rather self-selected. Furthermore, exposure to the package component in a network was determined by both the randomization scheme and visit attendance by index participants at visits when the initial intervention or boosters were delivered. A network was exposed if its index member attended a visit when a package component was administered (i.e., baseline, 6-month, and 12-month visits). Therefore, estimation of the effects of an intervention package component does not benefit from randomization, and adjustment for confounding is necessary to identify causal effects.

In studies with time-varying exposures subject to treatment confounder feedback, standard approaches to adjust for confounding, such as including time-varying confounders in an outcome model, are biased. Treatment confounder feedback occurs when a time-varying covariate is a risk factor for the outcome and affected by previous treatment because such a covariate is both a confounder and an intermediate variable [2, 3]. Adjusting for intermediate variables may result in conditioning on a collider which can lead to selection bias, while not adjusting for a confounder can result in confounding bias [4]. Instead, these time-varying confounders can be adjusted for using inverse probability weights, defined as the probability of component exposure conditional on past network-level exposure, network-level aggregate covariate history, and index member covariate history.

We adjust for possible confounding of index status by including baseline covariates in the outcome model. We define a model for the probability of exposure to the intervention package component at each visit, and a separate model is fit for each intervention component [5] conditional on the other component exposure histories, which is estimated using a logistic regression model in the intervention networks only pooled across study visits. This probability of exposure can be written as $\text{logit}(\Pr[A_{hkj} = a_{hkj} | \bar{\mathbf{A}}_{k,j-1}, \tilde{\mathbf{Z}}_{kij}, \bar{\mathbf{Z}}_{ki_kj}]) = \eta_0(j) + h(\bar{\mathbf{A}}_{k,j-1})\boldsymbol{\eta}_1 + h(\tilde{\mathbf{Z}}_{k,j-1})\boldsymbol{\eta}_2 + h(\bar{\mathbf{Z}}_{ki_k,j-1})\boldsymbol{\eta}_3$, which includes a time-dependent intercept $\eta_0(j)$. Define $h(\cdot)$ to be a known function of the treatment or covariate history with the dimensions of the column vectors $\boldsymbol{\eta}_1, \boldsymbol{\eta}_2, \boldsymbol{\eta}_3$ determined by $h(\cdot)$. Among the intervention networks with at least some network members observed in the study at visit $j$, the probability of exposure to component $h$ in network $k$ at visit $j$, with exposure denoted by $A_{hkj}$, conditional on prior exposure and covariate histories up to and including visit $j$ is defined as

$$\begin{aligned}\pi(\bar{\mathbf{A}}_{kj}, \bar{\mathbf{Z}}_{kij}, \boldsymbol{\eta}) &= f(A_{hkj}|\bar{\mathbf{A}}_{k,j-1}, \tilde{\mathbf{Z}}_{k,j-1}, \bar{\mathbf{Z}}_{ki_k,j-1}) \\ &= \frac{\exp(\eta_0(j) + h(\bar{\mathbf{A}}_{k,j-1})\boldsymbol{\eta}_1 + h(\tilde{\mathbf{Z}}_{k,j-1})\boldsymbol{\eta}_2 + h(\bar{\mathbf{Z}}_{ki_k,j-1})\boldsymbol{\eta}_3)}{1 + \exp(\eta_0(j) + h(\bar{\mathbf{A}}_{k,j-1})\boldsymbol{\eta}_1 + h(\tilde{\mathbf{Z}}_{k,j-1})\boldsymbol{\eta}_2 + h(\bar{\mathbf{Z}}_{ki_k,j-1})\boldsymbol{\eta}_3)},\end{aligned}$$

where $\boldsymbol{\eta} = (\eta_0(j), \boldsymbol{\eta}_1, \boldsymbol{\eta}_2, \boldsymbol{\eta}_3)$ denotes the vector of parameters. This model can be estimated using logistic regression pooled across visits in the network-level data, then the weights up to and including visit $j$ are cumulatively multiplied to obtain the probability of the network-level exposure up to and including a given study visit. Baseline covariates can be used to define stabilized weights that can lead to improvements in efficiency. We employed the following stabilized exposure weight [2] for each network $k$ observed from visit 2 to $M_k$, where $M_k$ is the max number of visits for participants in intervention network $k$:

$$w_{hk}(\bar{\mathbf{A}}_{kj}, \bar{\mathbf{Z}}_{kij}, \boldsymbol{\eta}_{Wh}) = \prod_{j=2}^{M_k} \frac{f(A_{hkj}|\bar{\mathbf{A}}_{k,j-1}, \tilde{\mathbf{Z}}_{k0}, \mathbf{Z}_{ki_k0})}{f(A_{hkj}|\bar{\mathbf{A}}_{k,j-1}, \tilde{\mathbf{Z}}_{kj}, \bar{\mathbf{Z}}_{ki_kj}, \tilde{\mathbf{Z}}_{k0}, \mathbf{Z}_{ki_k0})}.$$

Because these stabilized weights allow for exchangeability within levels of the baseline covariates $\tilde{\mathbf{Z}}_{k0}$ and $\mathbf{Z}_{ki_k0}$, these baseline covariates must also be included as covariates in the outcome model. When considering multiple components in a single outcome model, we can estimate the probability of exposure at each visit to each component separately conditional on past exposure and covariate history. Then, akin to multiplying multiple exposure weights to estimate joint causal effects [5, 6], we can multiply together the weights for each of the components included in the model. For example, suppose we had an outcome with three time-varying component exposures. In that case, the stabilized weight could be defined as $w(j) = w_{1k}(\bar{\mathbf{A}}_{kj}, \bar{\mathbf{Z}}_{kij}, \boldsymbol{\eta}_{W1}) \times w_{2k}(\bar{\mathbf{A}}_{kj}, \bar{\mathbf{Z}}_{kij}, \boldsymbol{\eta}_{W2}) \times w_{3k}(\bar{\mathbf{A}}_{kj}, \bar{\mathbf{Z}}_{kij}, \boldsymbol{\eta}_{W3})$.

#### Appendix B

Approach for multiple package components with pairwise interactions

We consider an approach to quantify the effects of multiple package components and include all pairwise interactions between components in a single model. We may also be interested in a specific contrast of combinations of the components that comprise the intervention package. For HIV prevention interventions, we may want to quantify the effect of exposure to HIV testing, condom promotion, and treatment as prevention, compared to treatment as prevention only. For HIV educational interventions, we could be interested in the effect of training from peer education and incentives, compared to peer education training only. A participant could receive these multiple components at a single visit or across different visits. Instead of using the model in equation (1), an alternative way to parameterize the outcome model is as follows:

$$\begin{aligned}\log\{E[Y_{ki,j+1}(r,\bar{\boldsymbol{a}}_j)|\mathbf{Z}_{ki},b_{ki},b_k]\} &= \beta_0 + r\beta_1 + g(\bar{\boldsymbol{a}}_j)\boldsymbol{\beta}_2 + rg(\bar{\boldsymbol{a}}_j)\boldsymbol{\beta}_3 \\ &+ \sum_{l<l'}^{H} g(\bar{\boldsymbol{a}}_{lj})^T g(\bar{\boldsymbol{a}}_{l'j})\boldsymbol{\beta}_{4ll'} + r\sum_{l<l'}^{H} g(\bar{\boldsymbol{a}}_{lj})^T g(\bar{\boldsymbol{a}}_{l'j})\boldsymbol{\beta}_{5ll'} \\ &+ \mathbf{Z}_{ki}^T\boldsymbol{\beta}_6 + b_{ki} + b_k \ ,\end{aligned}$$

where $g(\cdot)$ is a known function of the package component history and the parameters $\boldsymbol{\beta}_2$, $\boldsymbol{\beta}_3$, $\boldsymbol{\beta}_{4ll'}$, $\boldsymbol{\beta}_{5ll'}$ are column vectors with dimensions determined by $g(\cdot)$. For example, in a model with three binary package components, we denote the potential outcomes indexed by the vector of intervention component exposures $\bar{\boldsymbol{a}}_j = (\bar{\boldsymbol{a}}_{1j}, \bar{\boldsymbol{a}}_{2j}, \bar{\boldsymbol{a}}_{3j})$ and index status $r$. Two random intercepts, $b_{ki} \sim N(0,\psi_1)$ and $b_k \sim N(0,\psi_2)$, are included for correlation within participants and within networks, respectively. We used a log link with a working binomial distribution for the outcomes with an independent correlation specified through the random effects to obtain a robust estimator of the variance. The potential outcomes of interest are $Y_{ki,j+1}(r,\bar{\boldsymbol{a}}_{1j},\bar{\boldsymbol{a}}_{2j},\bar{\boldsymbol{a}}_{3j})$. This model could be expressed as:

$$\begin{aligned}\log\{E[Y_{ki,j+1}(r,\bar{\boldsymbol{a}}_{1j},\bar{\boldsymbol{a}}_{2j},\bar{\boldsymbol{a}}_{3j})|\mathbf{Z}_{ki},b_{ki},b_k]\} &= \beta_0 + r\beta_1 + g(\bar{\boldsymbol{a}}_{1j})\boldsymbol{\beta}_2 + g(\bar{\boldsymbol{a}}_{2j})\boldsymbol{\beta}_3 + g(\bar{\boldsymbol{a}}_{3j})\boldsymbol{\beta}_4 \\ &+ rg(\bar{\boldsymbol{a}}_{1j})\boldsymbol{\beta}_5 + rg(\bar{\boldsymbol{a}}_{2j})\boldsymbol{\beta}_6 + rg(\bar{\boldsymbol{a}}_{3j})\boldsymbol{\beta}_7 \\ &+ g(\bar{\boldsymbol{a}}_{1j})^T g(\bar{\boldsymbol{a}}_{2j})\boldsymbol{\beta}_8 + g(\bar{\boldsymbol{a}}_{1j})^T g(\bar{\boldsymbol{a}}_{3j})\boldsymbol{\beta}_9 + g(\bar{\boldsymbol{a}}_{2j})^T g(\bar{\boldsymbol{a}}_{3j})\boldsymbol{\beta}_{10} \\ &+ rg(\bar{\boldsymbol{a}}_{1j})^T g(\bar{\boldsymbol{a}}_{2j})\boldsymbol{\beta}_{11} + rg(\bar{\boldsymbol{a}}_{1j})^T g(\bar{\boldsymbol{a}}_{3j})\boldsymbol{\beta}_{12} + rg(\bar{\boldsymbol{a}}_{2j})^T g(\bar{\boldsymbol{a}}_{3j})\boldsymbol{\beta}_{13} \\ &+ \mathbf{Z}_{ki}^T\boldsymbol{\beta}_{14} + b_{ki} + b_k \ .\end{aligned}$$

For example, the effects of the first package component are: $\widehat{RR}^{D}(\bar{\boldsymbol{a}}) = \exp\Bigg(g(\bar{\boldsymbol{a}}_{1j})\hat{\boldsymbol{\beta}}_5\Bigg)$ and $\widehat{RR}^{S}(\bar{\boldsymbol{a}}) = \exp\Bigg(g(\bar{\boldsymbol{a}}_{1j})\hat{\boldsymbol{\beta}}_2\Bigg)$. The combined effects of the first and second component are: $\widehat{RR}^{D}(\bar{\boldsymbol{a}}) = \exp\Bigg(g(\bar{\boldsymbol{a}}_{1j})\hat{\boldsymbol{\beta}}_5 + g(\bar{\boldsymbol{a}}_{2j})\hat{\boldsymbol{\beta}}_6 + g(\bar{\boldsymbol{a}}_{1j})^T g(\bar{\boldsymbol{a}}_{2j})\hat{\boldsymbol{\beta}}_{11}\Bigg)$ and $\widehat{RR}^{S}(\bar{\boldsymbol{a}}) = \exp\Bigg(g(\bar{\boldsymbol{a}}_{1j})\hat{\boldsymbol{\beta}}_2 + g(\bar{\boldsymbol{a}}_{2j})\hat{\boldsymbol{\beta}}_3 + g(\bar{\boldsymbol{a}}_{1j})^T g(\bar{\boldsymbol{a}}_{2j})\hat{\boldsymbol{\beta}}_8\Bigg)$.

### Appendix C

Covariate selection procedure for modeling in HPTN 037

We provide additional details on the modeling approach, including the covariate selection procedure and the distribution of the stabilized weights for the booster exposure. Ideally, we would like to include all covariates that were known or suspected risk factors; however, given the size of our data set ($n = 560$), this resulted in too many parameters due to the number of independent covariates (a total of 19 covariates) and the need to include variables both for the index as well as the network. We thus employed the variable selection procedure described below.

Among all study participants, we determined the measured covariates that were associated with the outcome in the study data. We first fit univariate (unweighted) longitudinal models for each individual-level baseline covariate with the outcome any report of risk behavior during follow-up. We then included all individual-level baseline covariates with P value $< 0.20$ from univariate models in a single model and then performed backward selection to retain the individual-level baseline covariates with P value $< 0.20$, while forcing in the covariate for the report of any injection risk behavior at the prior visit because a measure prior behavior is a known important risk factor for subsequent outcomes. We included these selected individual-level baseline covariates in the unweighted outcome models and then selected baseline index member and network-level covariates in the inverse probability weighted (IPW) outcome model.

Because the time-varying (TV) booster exposure was not randomized, we need to adjust for TV index member covariates and TV network-level average covariates. As above, we determined the measured covariates that were associated with the outcome in the complete study data (both intervention and control networks with follow-up visits). We retained the index member and network-level report of any injection risk behavior outcome at the prior visit $j - 1$ as a covariate in the weight models for the exposure at visit $j$. Due to the use of stabilized weights, we included the baseline covariates in the numerator and denominator weight models and included these same baseline covariates in the outcome model. We also included pairwise interaction terms between the TV index and network-level covariates to fit our weight models more flexibly. The steps to determine the set of covariates for the outcome models and the estimation of the weights for the IPW models were as follows:

1. Fit separate univariate models for each index member baseline covariate with the outcome report of any risk behavior.

2. Include all index member baseline covariates with P value $< 0.20$ from the univariate model in the same outcome model and then perform backward selection, retain the index member baseline covariates with P value $< 0.20$.

3. In the outcome model, replace selected index member baseline covariates with their corresponding TV index member version, if applicable, regardless of the P value of the TV version of the variable in the outcome model.

4. Fit univariate models for each network-level baseline covariate with the outcome report of any risk behavior.

5. Add all network-level baseline covariates with P value $< 0.20$ from the univariate models to the model with the set of index member baseline and TV covariates selected in steps 2 and 3 and perform backward selection retaining only the network-level baseline covariates with P value $< 0.20$, while retaining the previously selected index member baseline and TV covariates.

6. In the outcome model, replace selected network-level baseline covariates with their corresponding TV network-level version, if applicable.

7. Generate all pairwise interaction terms between the selected TV index member and network-level covariates.

8. Add all the interaction terms generated to the model selected in step 6 and perform backward selection to keep only interaction terms with P value $< 0.20$ while retaining the selected baseline and TV index member and network-level covariates.

9. Include these selected baseline and TV index member and network-level covariates and selected interactions between TV index member and TV network-level covariates in the model for the denominator of the weights and include only the selected baseline index member and baseline network-level covariates in the model for numerator weights. These baseline index member and network-level covariates are also included in the fitted outcome model.

The time-varying status for index member injection of heroin and cocaine (and interactions involving that variable) was selected in the process, but not included in the weight models because of convergence issues. The unweighted outcome model was adjusted for baseline covariates only, then separately, their TV versions were included to estimate the unweighted outcome model adjusted for TV covariates. To note, because of convergence issues when fitting the outcome models adjusted for baseline and time-varying covariates to estimate the overall effects and other effects of the components, we had to replace the time-varying network-level covariate for prevalence of reporting any injection risk behaviors in the last month at the prior visit with its baseline version in these models. The lists of the selected covariates are included in each of these models is included in the footnotes of Table 3.

### Appendix D

#### Additional Results in HPTN 037

We provide details on how the notation in the main paper is linked to the application in HPTN 037. In this illustrative example, we estimated effects of the initial intervention denoted by $A_{1k0}$ and effects of the boosters denoted by $A_{2kj}$ across all study visits. The package components were defined as initial only and recent exposure to the 6- or 12-month booster in addition to the initial intervention. These effects describe the patterns of change in the probability of response over time in the study population. We defined a simple time-updated exposure for the booster variable. Assuming as in the primary analysis for this study that the initial intervention impacts study outcomes for the duration of follow-up and there is a baseline visit with five follow-up visits [7], the strategy "intended network intervention" is represented by $\bar{\boldsymbol{a}}_j = \begin{pmatrix} 1 & 1 & 1 & 1 & 1 & 1 \\ 0 & 1 & 1 & 0 & 0 & 0 \end{pmatrix}$ and the strategy "never exposed" is represented by $\bar{\mathbf{0}} = \begin{pmatrix} 0 & 0 & 0 & 0 & 0 & 0 \\ 0 & 0 & 0 & 0 & 0 & 0 \end{pmatrix}$. The observed network-level exposure for network $k$ up to visit $j$ is denoted by $\bar{\boldsymbol{A}}_{kj} = \begin{pmatrix} A_{1k0} & A_{1k0} & A_{1k0} & A_{1k0} & A_{1k0} & A_{1k0} \\ A_{2k0} & A_{2k1} & A_{2k2} & A_{2k3} & A_{2k4} & A_{2k5} \end{pmatrix}$.

Let $a_{10}$ denote a realization of the initial intervention randomized at baseline and $a_{2j}$ denote a realization of the package component for the boosters at visit $j$. The *direct* booster effect is a contrast in average potential outcomes under index versus network member status if the network is exposed to the initial and boosters; that is, on the ratio scale, $RR^D(a_{10}, a_{2j}) = E[Y_{ki,j+1}(r = 1, a_{10} = 1, a_{2j} = 1)]/E[Y_{ki,j+1}(r = 0, a_{10} = 1, a_{2j} = 1)]$. The *spillover* package effect compares the average potential outcomes if a participant is a network member under network exposure to the initial and booster versus no component history exposure $\bar{\mathbf{0}}$ (i.e., if the network is assigned to control); that is, $RR^S(a_{10}, a_{2j}) = E[Y_{ki,j+1}(r = 0, a_{10} = 1, a_{2j} = 1)]/E[Y_{ki,j+1}(r = 0, a_{10} = 0, a_{2j} = 0)]$. The *composite* package effect is $RR^{Comp}(a_{10}, a_{2j}) = E[Y_{ki,j+1}(r = 1, a_{10} = 1, a_{2j} = 1)]/E[Y_{ki,j+1}(r = 0, a_{10} = 0, a_{2j} = 0)]$; that is, a comparison of the average potential outcomes if an index under network component history exposure to initial and booster versus if a network member under no component history exposure $\bar{\mathbf{0}}$ (i.e., control network). Marginalizing over the index status, the *overall* package effect compares average potential outcomes under initial and booster exposure $\bar{\boldsymbol{a}}_j$ versus no exposure to either the initial or booster (i.e., control), denoted as $RR^{Overall}(a_{10}, a_{2j}) = E[Y_{ki,j+1}(a_{10} = 1, a_{2j} =$

$1)]/E[Y_{ki,j+1}(a_{10}=0, a_{2j}=0)]$[8]. The effects for the initial component alone can be defined similarly with $a_{10}=1, a_{2j}=0$.

The outcome model fit in the HPTN 037 data has separate terms for the initial intervention and the time-updated package intervention because these are components of the intervention package:

$$\log\{E[Y_{ki,j+1}(r, a_{10}, a_{2j})|\mathbf{Z}_{ki}, b_{ki}, b_k]\} = \beta_0 + \beta_1 r + \beta_2 a_{10} + \beta_3 a_{2j} + \beta_4 r a_{10} + \beta_5 r a_{2j} + \boldsymbol{\beta}_6 \mathbf{Z}_{ki}^T + b_{ki} + b_k \ .$$

Two random intercepts, $b_{ki} \sim N(0, \psi_1)$ and $b_k \sim N(0, \psi_2)$, are included for correlation within participants and within networks, respectively. We used a log link with a working binomial distribution for the outcomes with an independent correlation specified through the random effects to obtain a robust estimator of the variance. In this specific model, possible estimators for each of the parameters for the booster effects in the presence of the initial intervention are:

$$\begin{aligned}
\widehat{RR}^{D}(a_{10}, a_{2j}) &= \Big(\widehat{E}[Y_{ki,j+1}(r=1, a_{10}=1, a_{2j}=1)]/\widehat{E}[Y_{ki,j+1}(r=0, a_{10}=1, a_{2j}=1)]\Big)\exp(\hat{\beta}_1)^{-1} \\
&= \exp(\hat{\beta}_4 + \hat{\beta}_5) \\
\widehat{RR}^{S}(a_{10}, a_{2j}) &= \widehat{E}[Y_{ki,j+1}(r=0, a_{10}=1, a_{2j}=1)]/\widehat{E}[Y_{ki,j+1}(r=0, a_{10}=0, a_{2j}=0)] \\
&= \exp(\hat{\beta}_2 + \hat{\beta}_3) \\
\widehat{RR}^{Comp}(a_{10}, a_{2j}) &= \Big(\widehat{E}[Y_{ki,j+1}(r=1, a_{10}=1, a_{2j}=1)]/\widehat{E}[Y_{ki,j+1}(r=0, a_{10}=0, a_{2j}=0)]\Big)\exp(\hat{\beta}_1)^{-1} \\
&= \exp\left(\hat{\beta}_2 + \hat{\beta}_3 + \hat{\beta}_4 + \hat{\beta}_5\right) \ .
\end{aligned}$$

In HPTN 037, the initial intervention was randomized, and the component exposure status depended on the covariates. Exposure to the booster was defined as a time-updated variable for the booster exposure at time $j$ with the outcome at time $j+1$; therefore, the weights at baseline were one and the stabilized weights at the six-month and twelve-month visits also equaled one for both intervention and control networks because the numerator was the same as the denominator. Furthermore, because we assumed that networks randomized to the control could not later be exposed to intervention components, the non-randomized component exposure only occurred in the intervention group. Therefore, the weights were estimated from the intervention network data using records from the 12- and 18-month visits only, corresponding to component exposures at the 6- and 12-month visits. The stabilized weights were set to one for the control networks for all visits and for the intervention networks for the six-month and 12-month visits, and weights at the 24-month or later visits were the same as the 18-month visit.

Because stabilized weights were used in the estimation procedure for the MSMs, we evaluated if the mean of the stabilized weights was one. Departures from this weight distribution could indicate a violation of positivity and/or model misspecification [9]. Table A1 displays the distribution of the stabilized weights employed in the MSMs. The mean of the stabilized weights for time-varying exposure was approximately 1 across all study visits (and 0.98 at the 18-month visit only), providing no evidence of model misspecification [10] or possible violations of positivity (i.e., there is a positive probability of both exposure and no exposure for all possible combinations of the measured confounders in the study population). We also checked for random positivity violations via inspection of the cross-tabulation of each covariate with the peer education booster variable; none were noted. The calculations of the weights are described in Appendix A.

For the model used in this paper, a test of the $\beta_1$ term could provide information on the presence of common causes of index status and outcome. In the outcome model fit in the HPTN 037 data, the estimated bias correction term $\hat{\beta}_1$ for unmeasured confounding was rate ratio (RR) = 1.16 (95% confidence interval (CI) = 0.92, 1.48). Among the 560 participants, one isolate (participant with no others in their network in this analytic sample) was included in the analysis to improve estimation of the mixed models. Four person-visits had a missing value for one of the risk behaviors used to define the outcome. We coded these person-visits as no risk behavior reported.

In a sensitivity analysis, we evaluated effect measure modification by study visit on the multiplicative scale and estimated the effects at the 6-month visit (following the delivery of the baseline intervention) and 12- and 18-month visits (following the delivery of the booster intervention) using a MSM fit with inverse probability weights. At the six-month visit, the only relevant package component was the initial component delivered at baseline, so no estimates are reported for the booster at six months. We reported robust F-tests that tested the interaction term between each of the effects and visit month. Ignoring the estimation of the weights [11], we used a conservative empirical sandwich estimator of the variance from a generalized linear mixed model with a working binomial variance and independent correlation specified by within-participant and within-network random intercepts to construct 95% Wald-type CIs [12, 13]. The effect measure modification by study visit on the multiplicative scale for the four different effects is reported in Table A3. For the spillover effect of the boosters with initial, the magnitude of the effect was larger at 12 months (RR = 0.54; 95% confidence interval (CI) = (0.36, 0.81)), compared to 18 months (RR = 0.69; 95% CI = 0.44, 1.10). The magnitude of the estimated spillover effect of the initial intervention was comparable across study visits. The estimated direct effect of the initial intervention was null at six months and protective at 12 and 18 months. The estimated direct effect of the booster was protective at 18 months (RR = 0.86; 95% CI = 0.47, 1.58). The estimated composite effects of the initial and booster components were protective and the magnitude of the estimate for the initial component was larger at 18 months (RR = 0.57; 95% CI = 0.36, 0.91), compared to 12 months (RR = 0.70; 95% CI = 0.51, 0.98). The estimated composite effect of the booster was comparable at the 12 and 18 month visits (12-month RR = 0.63; 95% CI = 0.41, 0.99; 18-month RR = 0.60; 95% CI = 0.36, 1.00). The estimated overall effect for the initial intervention was larger in magnitude for later visits (18-month RR = 0.77; 95% CI = 0.60, 0.99, compared to 13% reduction in the overall rate at six months (RR 0.87; 95% CI = 0.71, 1.06)). The estimated overall effect of the booster was somewhat attenuated at the 18-month visit (RR = 0.66; 95% CI = 0.45, 0.96), compared to the 12-month visit (RR = 0.58; 95% CI = 0.45, 0.96). We may have been underpowered because HPTN 037 was not designed to detect effect measure modification by study visit.

Table A1: Distribution of the stabilized weights employed in the inverse probability weighted models in HPTN 037 for the time-varying package component, booster exposure, in relation to report of any risk behavior outcome among 560 participants with 1,598 follow-up visits across all study visits

| **Weights** | Sum | Mean | SD | Median | 1% | 99% | Min | Max |
|---|---|---|---|---|---|---|---|---|
| Unstabilized | 2,009.5 | 1.257 | 2.664 | 1.000 | 1.000 | 4.581 | 1.000 | 59.35 |
| Stabilized | 1,582.6 | 0.990 | 0.079 | 1.000 | 0.810 | 1.102 | 0.101 | 2.133 |

Standard deviation (SD)

Table A2: Distribution of the stabilized weights employed in the inverse probability weighted models in HPTN 037 for the time-varying package component, booster exposure, in relation to report of any risk behavior outcome among 560 participants with 1,598 follow-up visits at the 18-month visit

| **Weights** | Sum | Mean | SD | Median | 1% | 99% | Min | Max |
|---|---|---|---|---|---|---|---|---|
| Unstabilized | 497.1 | 1.516 | 4.391 | 1.007 | 1.000 | 4.581 | 1.000 | 59.35 |
| Stabilized | 319.8 | 0.975 | 0.136 | 1.000 | 0.132 | 1.488 | 0.101 | 2.133 |

Standard deviation (SD)

Table A3: Multiplicative effect measure modification of estimated rate ratios (RR) using Marginal Structural Models fit with inverse probability weights for the direct, spillover, composite and overall effects of the HPTN 037 peer education package components on reducing report of any injection risk behavior (per person-visit) during follow-up with 95% confidence intervals (CI) among participants with at least one follow-up visit, by study visit (months)[1–3]

| | **Direct** | | | | **Spillover** | | | |
|---|---|---|---|---|---|---|---|---|
| | Initial | P-value[4] | Booster | P-value[4] | Initial | P-value[4] | Booster | P-value[4] |
| 6 months | 1.00 (0.68, 1.48) | 0.07 | NA[5] | 0.09 | 0.87 (0.67, 1.12) | 0.96 | NA[5] | 0.54 |
| 12 months | 0.81 (0.54, 1.21) | | 1.17 (0.65, 2.09) | | 0.87 (0.69, 1.10) | | 0.54 (0.36, 0.81) | |
| 18 months | 0.65 (0.38, 1.12) | | 0.86 (0.47, 1.58) | | 0.87 (0.66, 1.16) | | 0.69 (0.44, 1.10) | |
| | **Overall** | | | | **Composite** | | | |
| | Initial | P-value[4] | Booster | P-value[4] | Initial | P-value[4] | Booster | P-value[4] |
| 6 months | 0.87 (0.71, 1.06) | 0.35 | NA[5] | 0.36 | 0.87 (0.64, 1.19) | 0.17 | NA[5] | 0.12 |
| 12 months | 0.82 (0.68, 0.99) | | 0.58 (0.43, 0.78) | | 0.70 (0.51, 0.98) | | 0.63 (0.41, 0.99) | |
| 18 months | 0.77 (0.60, 0.99) | | 0.66 (0.45, 0.96) | | 0.57 (0.36, 0.91) | | 0.60 (0.36, 1.00) | |

[1] Analysis included a total of 560 participants and 1,598 person-visits with 509 events total. One participant was excluded due to missing baseline information on spending the night on the street (in the past 6 months) and spending time in jail (in the past 6 months) at baseline. The package components were defined as initial only and recent exposure for the 6- or 12-month booster in addition to the initial intervention.

[2] Baseline covariates included individual-level race (nonwhite vs. white), Hispanic (yes vs. no), report of any injection risk behavior (yes vs. no), injected daily in the last month (yes vs. no), and alcohol use (got drunk vs. no), index member race (nonwhite vs. white), index report of any injection risk behavior (yes vs. no), index injected daily in the last month (yes vs. no), and index injected heroin and cocaine (yes vs. no) and network-level average age and network-level prevalence of nonwhite race, report of any injection risk behavior, cocaine use, and injected heroin and cocaine.

[3] Adjusted for weights estimated using the same index member and network-level covariates included in the baseline model, time-varying network-level prevalence of report of any injection risk behavior, cocaine use, and injected heroin and cocaine, the time-varying index report of any injection risk behavior and index injected daily in the last month, and four interaction terms between selected time-varying covariates: index report of any injection risk behavior and injected daily in the last month; network-level prevalence of injected heroin and cocaine and index report of any injection risk behavior; network-level prevalence of injected heroin and cocaine and index injected daily in the last month; and network-level prevalence of injected heroin and cocaine and report of any injection risk behavior.

[4] P-value for multiplicative effect measure modification obtained from a robust F-test.

[5] At the six-month visit, the only relevant package component was the initial component delivered at baseline, so no estimates are reported for the booster at six months.

#### Appendix E

```
/*EXAMPLE SAS CODE*/
/*PURPOSE: Obtain Direct, Spillover, Composite effect estimates for package components*/
/*Package components: initial intervention; boosters plus initial intervention*/
/*Using empirical sandwich estimator of variance*/
/*that accounts for correlation between participants within networks*/
/*and across visits within participants*/
/*PROC GLIMMIX, METHOD=RMPL: produces marginal estimates*/
/*with working independent correlation specified by random effects */
/*for participants within networks and visits within participants*/
/*EXAMPLE DATASET: ANALYDATA.SAS7BDAT*/
/* mynkid = Network ID */
/* uid = Subject ID */
/* Venum = Visit (months)*/
/* any risk = Any report of injection-related risk behavior*/
/* mytreat = Network-level intervention*/
/* myindex = Index status*/
/* prev_nPEboost = booster status at previous visit j-1 for network*/
/* lagprev_nPEboost = booster status two visits prior j-2 for network*/
/* basecov = Vector of baseline covariates for all participants*/
/* basecov_index = Vector of baseline covariates for indexes*/
/* basecov_network = Vector of baseline covariates averaged in a network*/
/* timecov_index = Vector of time-varying covariates for indexes*/
/* timecov_network = Vector of time-varying covariates averaged in a network*/
/***************************************************************/
options ps=60 ls=80 nodate pageno=1 nofmterr ;
dm log "clear;" continue; dm out "clear;" continue;

libname datalib 'C:\HPTN 037\sasdata';
/*load format library*/
options fmtsearch = (hptn.formats);
```

```
/*Create temporary SAS dataset*/

data analydata;
set hptn.analydata;
run;

/*Stabilized weight model*/
/*Denominator*/
/*all pairwise interactions between covariates could be added to this model*/
title "Denominator of stabilized weights";
proc logistic data=analydata  descending ;
model prev_nPEboost = lagprev_nPEboost visnum  basecov_index basecov_network
                                               timecov_index timecov_network/link=logit;
output out=model1_demon  predprobs=I;
run;

data model1_denom;
set model1_denom;
*output the probability of the exposure received conditional on the covariates;
     if prev_nPEboost =0 then pdenom=IP_0;
else if prev_nPEboost =1 then pdenom=IP_1;
run;


/*Numerator*/
/*all pairwise interactions between covariates could be added to this model*/
title "Numerator of stabilized weights";
proc logistic data=analydata descending ;
model prev_nPEboost= lagprev_nPEboost visnum
                     basecov_index basecov_network/link=logit;
output out=model2_num  predprobs=I;
```

```
run;

data model2_num;
set model2_num;
*output the probability of the exposure received conditional on the covariates;
     if prev_nPEboost =0 then pnum=IP_0;
else if prev_nPEboost =1 then pnum=IP_1;
run;

/*Sort the data sets with the estimated weights*/
proc sort data=model1_denom;
by mynkid visnum;
run;

proc sort data=model2_num;
by mynkid visnum;
run;

proc sort data=analydata;
by mynkid visnum uid;
run;

/*Combine into a single data set for the outcome model*/
data finaldata;
     merge analydata
           model1_denom (keep= mynkid visnum pdenom)
           model2_num (keep= mynkid visnum pnum);
        by mynkid visnum;
if first.mynkid then do;
fexpw=1; fexpw_n=1;
end;
retain fexpw fexpw_n;
```

```
/*stabilized weights*/
        expw=pnum/pdenom;
        /*multiple across all person-time observations for each subject*/
fexpw=expw*fexpw;

/*Unstabilized weights*/
expw_n=1/pdenom;
fexpw_n=expw_n*fexpw_n;
run;

/*Marginal Structural Model*/
/*Weighted with stabilized exposure weight*/
/*Model for Overall Effect Estimator*/
proc glimmix data=finaldata empirical method=RMPL;
weight fexpw;
class mynkid uid;
      model  anyrisk = prev_nPEboost  mytreat visit
                          basecov basecov_index basecov_network
                  /dist=log link=binomial s cl;
random intercept/ subject=mynkid;
random intercept/ subject=uid(mynkid);
   estimate 'Overall Initial Effect'   mytreat 1/exp cl;
   estimate 'Overall Booster Effect'   mytreat 1 prev_nPEboost_new 1/exp cl;
    run;

/*Model for Direct, Spillover, and Composite Effect Estimator*/
proc glimmix data=finaldata empirical method=RMPL;
weight fexpw;
class mynkid_n uid_n;
      model  anyrisk =  prev_nPEboost  mytreat visit
                          basecov basecov_index basecov_network
                          /dist=log link=binomial s cl;
```

```
random intercept/ subject=mynkid_n;
random intercept/ subject=uid(mynkid);
estimate 'Direct Initial Effect'     mytreat*myindex 1/exp cl;
estimate 'Spillover Initial Effect'  mytreat 1/exp cl;
estimate 'Composite Initial Effect'  mytreat*myindex 1 mytreat 1/exp cl;

estimate 'Direct Booster Effect'     mytreat*myindex 1 prev_nPEboost*myindex 1/exp cl;
estimate 'Spillover Booster Effect'  mytreat 1 prev_nPEboost 1/exp cl;
estimate 'Composite Booster Effect'  mytreat 1 mytreat*myindex 1
                                    prev_nPEboost 1 prev_nPEboost_new*myindex 1/exp cl;
run;
```

#### References

1. Hernán MA, Robins JM. Using big data to emulate a target trial when a randomized trial is not available. *American Journal of Epidemiology.* 2016; 183(8):758–764.

2. Hernán MÁ, Brumback B, Robins JM. Marginal structural models to estimate the causal effect of zidovudine on the survival of HIV-positive men. *Epidemiology.* 2000; 11(5):561–571.

3. Robins JM, Hernan MA, Brumback B. Marginal structural models and causal inference in epidemiology. *Epidemiology.* 2000; 11(5):550–560.

4. Cole SR, Platt RW, Schisterman EF, Chu H, Westreich D, Richardson D, et al. Illustrating bias due to conditioning on a collider. *International Journal of Epidemiology.* 2010; 39(2):417–420.

5. Howe CJ, Cole SR, Mehta SH, Kirk GD. Estimating the effects of multiple time-varying exposures using joint marginal structural models: alcohol consumption, injection drug use, and HIV acquisition. *Epidemiology.* 2012; 23(4):574–582.

6. Hernán MA, Brumback B, Robins JM. Marginal structural models to estimate the joint causal effect of nonrandomized treatments. *Journal of the American Statistical Association.* 2001; 96(454):440–448.

7. Latkin CA, Donnell D, Metzger D, Sherman S, Aramrattna A, Davis-Vogel A, et al. The efficacy of a network intervention to reduce HIV risk behaviors among drug users and risk partners in Chiang Mai, Thailand and Philadelphia, USA. *Social Science and Medicine.* 2009; 68(4):740–748.

8. Buchanan A, Vermund S, Friedman S, Spiegelman D. Assessing Individual and Disseminated Effects in Network-Randomized Studies. *American Journal of Epidemiology.* 2018; 187(11):2449–2459.

9. Cole SR, Hernán MA. Constructing inverse probability weights for marginal structural models. *American Journal of Epidemiology.* 2008; 168(6):656–664.

10. Bang H, Robins JM. Doubly robust estimation in missing data and causal inference models. *Biometrics.* 2005; 61(4):962–973.

11. Hernán MA, Brumback BA, Robins JM. Estimating the causal effect of zidovudine on CD4 count with a marginal structural model for repeated measures. *Statistics in Medicine.* 2002; 21 (12):1689–1709.

12. Fitzmaurice GM, Laird NM, Ware JH. *Applied Longitudinal Analysis*. Hoboken: John Wiley & Sons, 2012.

13. Stefanski LA, Boos DD. The calculus of M-estimation. *The American Statistician*. 2002; 56(1):29–38.
